# Development and Internal Validation of a County-Level Screening Index for Postpartum Medicaid Access Barriers

**DOI:** 10.64898/2026.07.05.26357332

**Authors:** Caroline Howard, Prashant Shekhar

**Affiliations:** Department of Mathematics, Embry-Riddle Aeronautical University, Daytona Beach, FL, USA

**Author notes:** **Correspondence:** Caroline Howard. **Declaration of Interest** The authors declare no competing interests.

**Keywords:** Medicaid, postpartum care, maternal health, health services research, access to care, rural health, administrative burden, county-level index, spatial analysis, policy screening

## Abstract

**Background:** Postpartum Medicaid coverage and support are central maternal health policy issues, but county-level tools for identifying where postpartum Medicaid populations may face overlapping administrative, clinical, and contextual access barriers remain limited.

**Methods:** We developed and internally validated a county-level Postpartum Medicaid Access Barrier Index for all 3,144 counties and county equivalents in the 50 states and District of Columbia. Public data sources included geocoded Medicaid office locations from Shafer et al. (2024), U.S. Census county boundaries, American Community Survey 2024 5-year county indicators, the National Center for Health Statistics 2023 Urban-Rural Classification Scheme for Counties, and county-level hospital-based obstetric care status from the University of Minnesota Rural Health Research Center. Medicaid office locations were spatially assigned to counties, then merged with ACS indicators, rurality, and obstetric care status by county FIPS. The theoretical score range was 0–11; the index assigned higher weights to two core infrastructure measures and lower weights to contextual indicators. Internal validation assessed component structure, known-groups validity, geographic clustering, weighting sensitivity, added value over simpler infrastructure screens, and separation across concern levels.

**Results:** Across 3,144 counties, observed scores ranged from 0 to 10 on the theoretical 0–11 score, with a mean of 3.65 and median of 3. High or highest concern counties accounted for 665 counties (21.2%), including 56 counties (1.8%) in the highest concern group. Component correlations were low-to-moderate, with an average absolute phi of 0.176 and no pairwise component correlation at or above 0.50. Known-groups validity was strong: dual administrative and clinical gap counties scored 4.43 points higher than counties with neither gap (Cohen’s *d* = 3.28, *p* < 0.001). Scores were geographically clustered (Moran’s *I* = 0.375, permutation *p* = 0.005). A dual-gap-only screen captured 386 of 665 high/highest concern counties (58.0%) but missed 279 high/highest counties; a parsimonious rule requiring one infrastructure gap plus at least four contextual flags recovered 265 of these 279 missed counties (95.0%) with 100.0% precision.

**Discussion:** The Postpartum Medicaid Access Barrier Index provides a transparent county-level screening tool for identifying places where administrative, clinical, and contextual barriers may overlap for postpartum Medicaid populations and should be externally validated against Medicaid enrollment, renewal, churn, coverage continuity, and postpartum care outcomes.

## Introduction

Medicaid is central to pregnancy-related coverage and postpartum health financing in the United States, and recent policy attention has focused on extending postpartum Medicaid coverage and improving continuity after delivery (Centers for Medicare & Medicaid Services, 2021; Kaiser Family Foundation, 2024). This focus reflects evidence that coverage around childbirth is often unstable for low-income women, with substantial insurance churn in the months before and after delivery (Daw et al., 2017).

Coverage continuity depends on more than eligibility policy. Postpartum Medicaid populations may also face administrative burden, transportation barriers, digital access constraints, language access needs, disability-related support needs, and geographically uneven maternity care infrastructure (Herd & Moynihan, 2018; Syed et al., 2013). Rural hospital obstetric services have declined over time, and loss of hospital-based obstetric care has been linked to worsening maternity care access in rural counties (Hung et al., 2017; Kozhimannil et al., 2018).

These domains are typically measured separately. Medicaid office locations describe in-person administrative infrastructure, ACS indicators describe county population context, rural-urban classifications describe geography, and hospital obstetric care status describes clinical maternity infrastructure. For screening and program-context review, the harder question is where these barriers overlap. Because individual postpartum Medicaid enrollment and claims outcomes were not available for this development study, the index was designed to identify county-level contexts where barriers relevant to postpartum Medicaid access may overlap, rather than to measure postpartum Medicaid access directly. This study develops and internally validates a county-level Postpartum Medicaid Access Barrier Index that integrates public administrative, clinical, demographic, and spatial data to support county and state comparison, access-support review, and component-level interpretation.

## Methods

### Study Design and Analytic Aim

We conducted a county-level index development and internal validation study. The unit of analysis was the county or county equivalent. The index was built around two prespecified domains: direct access infrastructure and contextual access-support burden. Direct infrastructure included Medicaid office availability as an administrative/enrollment support measure and hospital-based obstetric care availability as a local maternity care infrastructure measure. Contextual access-support burden included poverty, transportation access, digital access, language access, disability-related support context, reproductive-age demand context, and rurality.

### Analytic Universe

The analytic universe included all 3,144 counties and county equivalents in the 50 states and District of Columbia. Puerto Rico and other territories were excluded because the Medicaid office dataset and analytic framing were limited to the 50 states and District of Columbia. County FIPS codes were used as the primary linkage key across all county-level sources. FIPS codes were stored as five-character text fields to preserve leading zeros.

### Data Sources

#### Medicaid office locations

Medicaid office locations were obtained from the Shafer et al. (2024) geocoded Medicaid office locations dataset from Harvard Dataverse and used to identify whether a county had at least one Medicaid office. This measure captures in-person administrative or enrollment support infrastructure, but it does not capture all application, renewal, or enrollment assistance available through online systems, phone support, hospitals, navigators, community organizations, managed care organizations, neighboring counties, or other local agencies.

#### County geography

U.S. Census 2025 cartographic county boundaries `(cb_2025_us_county_500k)` were used to assign Medicaid office points to counties and to build a complete county reference table, retaining counties with zero Medicaid offices in the denominator.

#### ACS county indicators

ACS 2024 5-year county-level indicators were obtained from the U.S. Census API. Scoring and contextual indicators included poverty rate, no-vehicle household rate, no-internet subscription rate, limited-English rate, disability rate, and female population ages 15–44 rate. Female population ages 15–44 was included as a reproductive-age demand context measure, not as a direct access barrier or measure of postpartum Medicaid enrollment.

#### Rural-urban classification

Rural-urban context was added using the National Center for Health Statistics 2023 Urban-Rural Classification Scheme for Counties. Counties were classified into six NCHS categories and summarized into a metro/nonmetro flag.

#### Hospital-based obstetric care status

County-level hospital-based obstetric care status was obtained from the University of Minnesota Rural Health Research Center’s 2010–2024 County-Level Hospital-Based Obstetric Care Status dataset and used as the clinical maternity infrastructure measure. This measure captures local hospital-based obstetric care availability, not outpatient postpartum care, postpartum visit availability, provider capacity, Medicaid provider participation, behavioral health, lactation support, or other postpartum services.

### Data Processing and Linkage

Source layers were converted into a single county-level analytic file. Medicaid office records were cleaned, restricted to valid coordinates, spatially assigned to Census 2025 counties, summarized as county office counts, and merged to a complete county reference table. ACS indicators, NCHS rural-urban classification, and hospital-based obstetric care status were merged by county FIPS. ACS rates were calculated from raw counts, and top-quartile flags were calculated across the 50-state-plus-DC county universe.

### Index Construction

The index was designed as a formative screening index. Components were selected because they represent distinct access-barrier domains relevant to postpartum Medicaid access, and each component remains visible in the analytic file.

The theoretical 0–11 score used two point values. Two core infrastructure measures received 2 points each, and seven contextual access-support measures received 1 point each (Table 1).

**Table 1.** Index components and point values.

| Component | Points |
| --- | --- |
| *Direct infrastructure measures* |  |
| No Medicaid office in county | 2 |
| No hospital-based obstetric care | 2 |
| *Contextual access-support measures* |  |
| Top-quartile poverty rate | 1 |
| Top-quartile no-vehicle household rate | 1 |
| Top-quartile no-internet subscription rate | 1 |
| Top-quartile limited-English rate | 1 |
| Top-quartile disability rate | 1 |
| Top-quartile female population ages 15–44 rate | 1 |
| Nonmetro county | 1 |

Weights were specified before validation. Medicaid office absence and hospital-based obstetric care absence received 2 points each because they represent direct administrative and clinical infrastructure domains. Contextual measures received 1 point each because they identify conditions that may make infrastructure gaps harder to navigate or may indicate greater reproductive-age demand context. Quartile thresholds were selected to support transparent national screening and component comparability, not to represent clinical, statutory, or program eligibility cut points. Validation analyses then tested whether this specification was stable under equal-weight, admin/clinical-heavy, context-only, and component-exclusion alternatives. Disability rate was included as an access-support context measure; female population ages 15–44 was included as reproductive-age demand context.

Concern categories were defined over the theoretical score range (Table 2). Observed county scores ranged from 0 to 10; no county scored 11.

**Table 2.** Concern-level definitions.

| Score range | Concern level |
| --- | --- |
| 0–2 | Lower concern |
| 3–5 | Moderate concern |
| 6–8 | High concern |
| 9–11 | Highest concern |

**Table 3.** Score distribution and concern levels.

| Measure | Count / value | Percent |
| --- | --- | --- |
| Counties/county equivalents | 3,144 | 100.0 |
| Observed score range | 0–10 | – |
| Mean score | 3.65 | – |
| Median score | 3 | – |
| Lower concern | 1,070 | 34.0 |
| Moderate concern | 1,409 | 44.8 |
| High concern | 609 | 19.4 |
| Highest concern | 56 | 1.8 |
| High or highest concern | 665 | 21.2 |

**Table 4.**
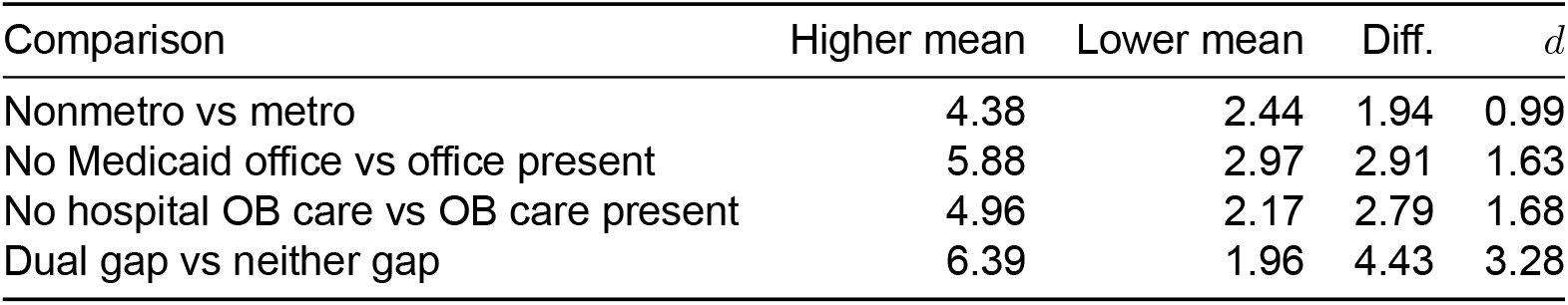
Known-groups validity comparisons.

| Comparison | Higher mean | Lower mean | Diff. | $d$ |
| --- | --- | --- | --- | --- |
| Nonmetro vs metro | 4.38 | 2.44 | 1.94 | 0.99 |
| No Medicaid office vs office present | 5.88 | 2.97 | 2.91 | 1.63 |
| No hospital OB care vs OB care present | 4.96 | 2.17 | 2.79 | 1.68 |
| Dual gap vs neither gap | 6.39 | 1.96 | 4.43 | 3.28 |

**Table 5.** Primary score drivers among high/highest concern counties.

| Driver group | Counties | Percent | Mean score |
| --- | --- | --- | --- |
| Dual infrastructure gaps | 386 | 58.0 | 7.21 |
| Clinical-only gap | 244 | 36.7 | 6.48 |
| Administrative-only gap | 21 | 3.2 | 6.62 |
| Contextual burden only | 14 | 2.1 | 6.00 |

**Table 6.**
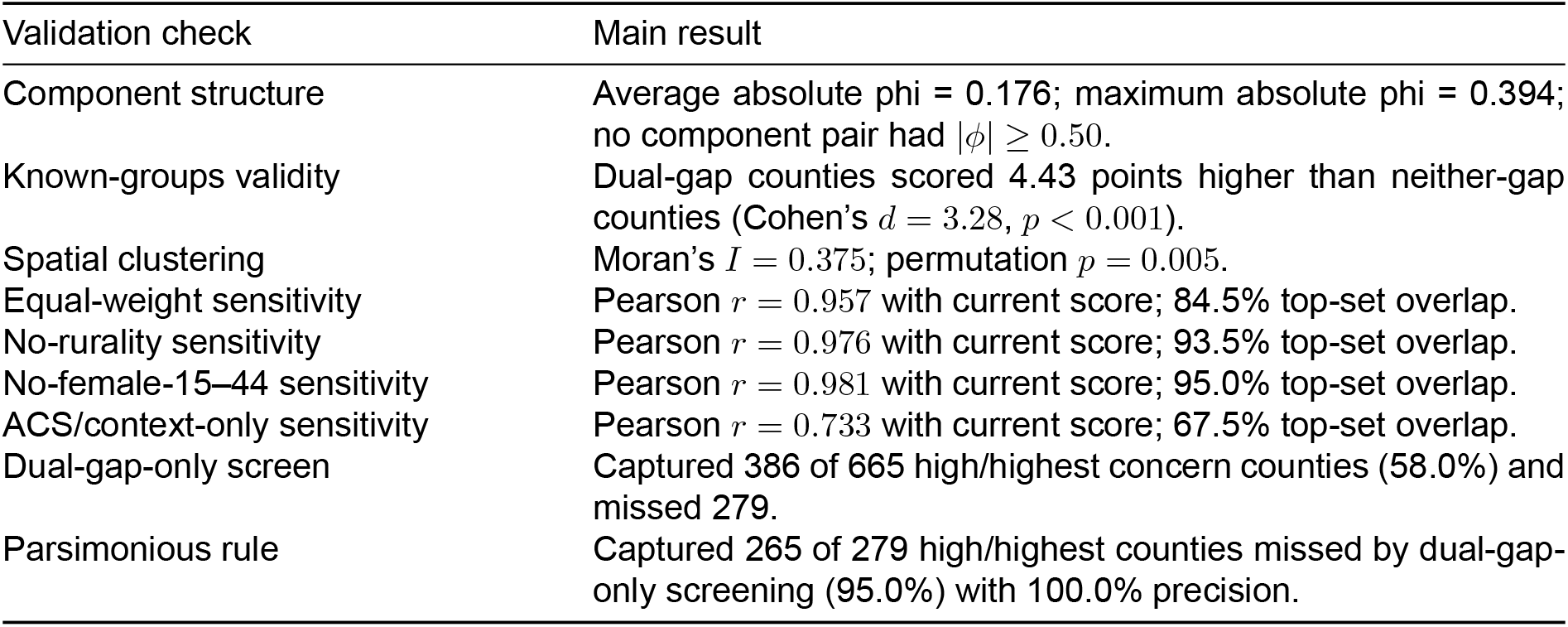
Sensitivity and added-value validation checks.

### Validation Framework

Internal validation focused on five questions: whether components were nonredundant, whether scores separated expected higher-burden groups, whether scores were geographically clustered, whether findings were stable under alternate weights, and whether the full score added information beyond simpler infrastructure screens. These analyses support construct validity, known-groups validity, robustness, and spatial patterning; they do not establish predictive or outcome validity. Component structure was assessed with pairwise phi correlations. Known-groups validity compared metro/nonmetro counties, counties with and without Medicaid offices, counties with and without hospital-based obstetric care, and dual-gap versus neither-gap counties. Spatial clustering was assessed using Moran’s *I* with 8-nearest-neighbor county-centroid weights and 199 permutations to provide a transparent county-neighbor structure while avoiding reliance on shared-border definitions that can be sensitive to county size and noncontiguous geographies. Weighting sensitivity compared the prespecified score with equal-weight, admin/clinical-heavy, ACS/context-only, no-rurality, and no-female-15–44 specifications. Added-value analyses compared the full high/highest concern set with Medicaid-office-only, obstetric-care-only, either-gap, and dual-gap-only screens. Kruskal-Wallis tests compared contextual indicators across concern levels.

## Results

### Score Distribution

Observed scores ranged from 0 to 10 on the theoretical 0–11 score. The mean score was 3.65, the median score was 3, and the population standard deviation was 2.17. Lower concern counties accounted for 1,070 counties, moderate concern for 1,409 counties, high concern for 609 counties, and highest concern for 56 counties. Overall, 665 counties (21.2%) were classified as high or highest concern.

Component prevalence varied across domains. No Medicaid office was present in 733 counties (23.3%), while no hospital-based obstetric care was present in 1,660 counties (52.8%). Nonmetro status applied to 1,958 counties (62.3%). By construction, each top-quartile ACS indicator applied to approximately 786 counties (25.0%).

### Component Structure

Component correlations were low-to-moderate. The average absolute phi correlation across component pairs was 0.176. The maximum absolute phi correlation was 0.394, and no component pair had an absolute correlation at or above 0.50. The strongest positive component relationships were high poverty with high no-internet rate (phi = 0.394), high poverty with high disability rate (phi = 0.383), high no-internet rate with high disability rate (phi = 0.347), high poverty with high no-vehicle rate (phi = 0.338), and no hospital-based obstetric care with high no-internet rate (phi = 0.337). These results indicate that components are related in theoretically plausible ways but are not redundant.

### Known-Groups Validity and Metro/Nonmetro Differences

Known-groups comparisons moved in the expected direction. Nonmetro counties had a higher mean score than metro counties (4.38 vs 2.44). Counties without a Medicaid office had a higher mean score than counties with a Medicaid office (5.88 vs 2.97). Counties without hospital-based obstetric care had a higher mean score than counties with hospital-based obstetric care (4.96 vs 2.17).

The strongest separation was observed for dual-gap counties. Counties with neither Medicaid office absence nor hospital-based obstetric care absence had a mean score of 1.96, while dual-gap counties had a mean score of 6.39. The mean difference was 4.43 points, with Cohen’s *d* = 3.28 and *p* < 0.001. Only 14 of 1,329 counties with neither gap (1.1%) were high/highest concern, compared with 386 of 578 dual-gap counties (66.8%).

Metro/nonmetro differences further supported known-groups validity. Among 1,958 nonmetro counties, 597 (30.5%) were high/highest concern. Among 1,186 metro counties, 68 (5.7%) were high/highest concern. Nonmetro counties also had higher prevalence of Medicaid office absence, hospital-based obstetric care absence, and dual-gap status.

### State Concentration and Spatial Clustering

High scores were concentrated in particular states and regions. The states with the highest average county scores were Mississippi (6.13), Louisiana (6.02), Alabama (5.67), New Mexico (5.52), Alaska (5.50), and Oklahoma (5.29). Texas had the largest count of high/highest concern counties, with 89 counties. Mississippi had the highest share of high/highest concern counties, with 54 of 82 counties (65.9%).

These results identify counties and states that may warrant closer review in future external validation and program-context analyses, especially Mississippi, Louisiana, Alabama, New Mexico, Oklahoma, Alaska, and Texas. Within those states, the index highlights specific counties where administrative infrastructure, clinical maternity infrastructure, and contextual access-support burden overlap.

The spatial clustering analysis supported this geographic interpretation. County scores were spatially clustered, with Moran’s *I* = 0.375 using 8-nearest-neighbor county-centroid weights. In 199 random permutations, the permutation *p* value was 0.005 and the mean permuted Moran’s *I* was 0.001, indicating that high and low scores were geographically patterned rather than randomly distributed across counties.

### Weighting Sensitivity

The current score was robust to reasonable alternate scoring specifications. The equal-weight score was highly correlated with the current score (Pearson *r* = 0.957; Spearman *r* = 0.951) and preserved 562 of the 665 high/highest concern counties (84.5%) in the same-size top set. The admin/clinical-heavy score was also highly correlated (Pearson *r* = 0.985; Spearman *r* = 0.984), preserving 579 counties (87.1%). Removing rurality yielded Pearson *r* = 0.976 and preserved 622 counties (93.5%). Removing female ages 15–44 yielded Pearson *r* = 0.981 and preserved 632 counties (95.0%).

The ACS/context-only specification showed lower agreement (Pearson *r* = 0.733; Spearman *r* = 0.699; top-set overlap = 449 counties, 67.5%). This was expected because that specification removes the two intentionally weighted core infrastructure measures. The result supports the interpretation that the full index is not reducible to demographic or socioeconomic context alone; it captures the intended overlap between administrative infrastructure, clinical maternity infrastructure, and contextual access-support burden.

### Infrastructure-Stratified Access-Support Burden

The contextual components differentiated counties within infrastructure groups. Counties with neither infrastructure gap had an average support-burden score of 1.96; admin-only gap counties had an average support-burden score of 1.96; clinical-only gap counties had an average support-burden score of 2.20; and dual-gap counties had an average support-burden score of 2.39. High access-support burden, defined as support score ≥ 4, appeared in 160 of 1,329 counties with neither gap (12.0%), 21 of 155 admin-only counties (13.5%), 244 of 1,082 clinical-only counties (22.6%), and 150 of 578 dual-gap counties (26.0%).

Among dual-gap counties, 150 of 578 (26.0%) also had high access-support burden, indicating counties that combined no Medicaid office, no hospital-based obstetric care, and at least four contextual indicators. This supports a layered interpretation: infrastructure components identify direct administrative and clinical infrastructure gaps, while ACS/rurality components identify where those gaps may be harder to navigate and therefore warrant closer review.

For practical screening, dual-gap counties with high contextual access-support burden represent a key hypothesis-generating group. A second review group is clinical-only gap counties with high support burden, because they may lack local hospital-based obstetric care while also having elevated poverty, transportation, digital, disability, language, reproductive-age demand, or rurality-related context. A third review group is the smaller set of high-contextual-burden counties without both infrastructure gaps, which may warrant further review despite appearing lower risk under infrastructure-only screens.

### Score-Driver Classification Among High-Concern Counties

We classified high/highest concern counties by the infrastructure and contextual components driving their scores. Among 665 high/highest concern counties, 386 (58.0%) had dual infrastructure gaps, meaning they had both no Medicaid office and no hospital-based obstetric care. Another 244 counties (36.7%) had a clinical-only gap with high contextual access-support burden. Twenty-one counties (3.2%) had an administrative-only gap with high contextual access-support burden, and 14 counties (2.1%) reached high/highest concern through contextual burden without either direct infrastructure gap.

This classification clarifies the county review sequence produced by the index. The largest high-concern group was dual-gap counties, but 279 high/highest concern counties were not dual-gap counties. Nearly all of these non-dual-gap counties reflected one infrastructure gap plus high contextual burden, especially clinical-only gaps. In the highlighted states, high/highest concern counties were often dual-gap counties: 43 of 54 in Mississippi, 29 of 36 in Louisiana, 35 of 37 in Alabama, 35 of 40 in Oklahoma, 17 of 18 in Alaska, and 70 of 89 in Texas. New Mexico showed a more mixed pattern, with 6 dual-gap, 5 clinical-only, 4 administrative-only, and 1 contextual-burden-only high/highest concern county.

### Applied County-Priority Interpretation

The score-driver classification was translated into a compact two-tier screening interpretation. The first tier identifies counties with direct infrastructure gaps; the second tier uses contextual access-support burden to distinguish which infrastructure-gap counties may warrant closer hypothesis-generating review. The 11 maximum-score counties all combined both infrastructure gaps, non-metro status, high poverty, high no-vehicle household rate, high no-internet subscription rate, and high disability rate. Detailed county lists and follow-up group summaries are provided in the supplement.

### Added Value Over Infrastructure-Only Screening

The full index identified high/highest concern counties that simpler infrastructure screens would miss. A Medicaid-office-only screen captured 407 of 665 high/highest concern counties (61.2%) but missed 258 counties. An obstetric-care-only screen captured 630 of 665 counties (94.7%) but missed 35 counties. An either-infrastructure-gap screen captured 651 of 665 counties (97.9%) but missed 14 counties with high contextual burden despite no direct infrastructure gap. A dual-gap-only screen captured 386 of 665 high/highest counties (58.0%) but missed 279 high/highest counties.

High/highest counties missed by dual-gap-only screening had an average score of 6.46 and an average support-burden score of 4.56. Most were nonmetro (95.3%) and had high poverty (90.0%), high no-internet rates (83.5%), high disability rates (75.6%), or high no-vehicle rates (71.0%).

### Parsimonious Rule Test

We tested whether a simpler contextual rule could recover high/highest counties missed by dual-gap-only screening. The target set included 279 high/highest concern counties not captured by dual-gap-only screening. A rule requiring one infrastructure gap plus at least four broad contextual flags captured 265 of these 279 counties (95.0%) with 100.0% precision and no false positives.

### Concern-Level Separation

Kruskal-Wallis tests showed significant differences across concern levels for all tested contextual indicators (all *p* < 0.001). Median poverty rates increased from 11.4% in lower concern counties to 24.3% in highest concern counties. Median no-vehicle rates increased from 5.0% to 9.2%, median no-internet rates from 7.6% to 19.1%, and median disability rates from 14.5% to 22.1%. The access-support burden score showed clear ordered separation, with median scores of 1, 2, 4, and 5 across lower, moderate, high, and highest concern counties, respectively. Limited-English rate and female ages 15–44 rate also differed across concern levels, but these patterns were less monotonic; female ages 15–44 should be interpreted as reproductive-age demand context rather than an access barrier.

## Discussion

The index identifies a clear geography for further review and external validation. High scores were concentrated in the South, parts of the Southwest, and Alaska. Mississippi, Louisiana, Alabama, New Mexico, Alaska, Oklahoma, and Texas may warrant closer review in future validation and program-context analyses because they represent different screening patterns: broad county-level burden, large absolute numbers of high/highest concern counties, rurality and distance, or mixed score-driver profiles.

The applied priority interpretation clarifies how the index can be used as a screening tool. The 11 maximum-score counties represent the clearest hypothesis-generating case-review group because every one combines no Medicaid office, no hospital-based obstetric care, nonmetro status, high poverty, high no-vehicle rate, high no-internet rate, and high disability rate. They represent the most concentrated version of the barrier profile the index was designed to identify.

The two-tier framework then separates counties by potential follow-up question. Dual-gap counties with high contextual burden point to places where administrative access planning and maternity care access planning may need to be considered together. One-gap counties with high contextual burden point to a different problem: they may not look like the most severe counties under an infrastructure-only screen, but the remaining infrastructure gap overlaps with multiple socioeconomic, transportation, digital, disability, language, reproductive-age demand, or rurality-related indicators. These counties are central to the index’s contribution because they are the counties most likely to be missed by a dual-gap-only approach.

The added-value and parsimonious-rule analyses clarify this contribution. Counties missed by dual-gap-only screening were not low-burden outliers; they had substantial contextual access-support burden. The parsimonious rule showed that much of this missed-county signal can be summarized as one infrastructure gap plus at least four contextual flags. In applied use, those counties may warrant examination alongside dual-gap counties rather than being excluded by a narrower infrastructure-only screen. The full index adds value because it preserves component detail, distinguishes concern levels, supports state comparison, and retains the smaller contextual-burden watchlist.

### Policy and Program Implications

The index can support hypothesis generation and program-context review by turning national county-level data into a sequence of reviewable county groups. As an additional contribution, the project includes a Power BI visual analysis tool that allows users to examine the data directly through county maps, ranked county lists, state comparisons, and component-level score drivers. This visual layer supports exploratory analysis of which counties are high scoring, which components drive those scores, and how patterns differ across states or metro/nonmetro contexts.

Future work should link the index to Medicaid enrollment, renewal, churn, postpartum coverage continuity, and claims-based postpartum care data to test whether flagged counties show measurable program access or continuity concerns. This would determine whether the screening geography identified here translates into observed Medicaid administrative outcomes and would allow future versions of the index to be refined using linked program data.

### Limitations

Several limitations should guide interpretation. The index is county-level and may mask within-county variation in transportation networks, internet access, administrative assistance, and maternity care availability. County-level context also creates ecological inference risk: the analysis cannot show that postpartum Medicaid enrollees themselves experience the measured county-level barriers. The index does not identify individual postpartum Medicaid enrollees, pregnancy status, births, or postpartum coverage status; female population ages 15–44 is a reproductive-age demand context measure. Medicaid office availability is a noisy proxy for administrative access because states organize eligibility, enrollment, local offices, online assistance, phone assistance, navigators, managed care supports, and county social service functions differently. A county with no Medicaid office may still have meaningful enrollment assistance, and a county with an office may still have limited access because of distance, staffing, hours, language access, or appointment availability. Hospital-based obstetric care status does not capture outpatient obstetric care, postpartum care quality, provider capacity, Medicaid provider participation, behavioral health, lactation support, birth centers, or distance to care. ACS estimates are survey-based, and top-quartile thresholds indicate relative county burden rather than clinical or statutory cut points. Source years also differ across inputs, including ACS 2024, NCHS 2023 rurality, 2010–2024 hospital-based obstetric care status, 2024 Medicaid office data, and Census 2025 county boundaries. The internal validation analyses support construct validity, known-groups validity, robustness, and spatial patterning; external validation against Medicaid administrative outcomes remains the next step.

## Conclusions

The Postpartum Medicaid Access Barrier Index integrates Medicaid office availability, hospital-based obstetric care status, ACS access-support indicators, rurality, and reproductive-age demand context into a transparent county-level screening score. Internal validation supports its use for identifying counties where administrative, clinical, and contextual barriers may overlap for postpartum Medicaid populations. The index adds value over single-domain screening by identifying high-burden counties missed by Medicaid-office-only or dual-gap-only approaches and provides a foundation for future validation against Medicaid enrollment, renewal, churn, postpartum coverage continuity, and postpartum care utilization outcomes.

## Data Availability

Data availability: This study uses publicly available county-level data. Processed analytic files and documentation are available in the project repository: https://github.com/caroline-howard/postpartum-medicaid-access-barrier-index.

## Declarations

### Ethics approval

This study used publicly available county-level and facility/location data and did not involve human subjects or individual-level protected health information.

### Funding

No external funding was used for this draft.

### Competing interests

The authors declare no competing interests.

### Data availability

This study uses publicly available county-level data. Processed analytic files and documentation are available in the project repository: https://github.com/caroline-howard/postpartum-medicaid-access-barrier-index.

### Code availability

Analysis code is available in the project repository: https://github.com/caroline-howard/postpartum-medicaid-access-barrier-index.

## Supplementary Appendix

**Supplemental Table S1.** Maximum-Score Counties. The 11 counties with the maximum observed score of 10 all had both direct infrastructure gaps, nonmetro status, high poverty, high no-vehicle household rate, high no-internet subscription rate, and high disability rate.

| County | State | Score |
| --- | --- | --- |
| Barbour County | Alabama | 10 |
| Perry County | Alabama | 10 |
| Sumter County | Alabama | 10 |
| Evangeline Parish | Louisiana | 10 |
| Madison Parish | Louisiana | 10 |
| Humphreys County | Mississippi | 10 |
| Noxubee County | Mississippi | 10 |
| Harding County | New Mexico | 10 |
| Harmon County | Oklahoma | 10 |
| Tillman County | Oklahoma | 10 |
| Duval County | Texas | 10 |

**Supplemental Table S2.**
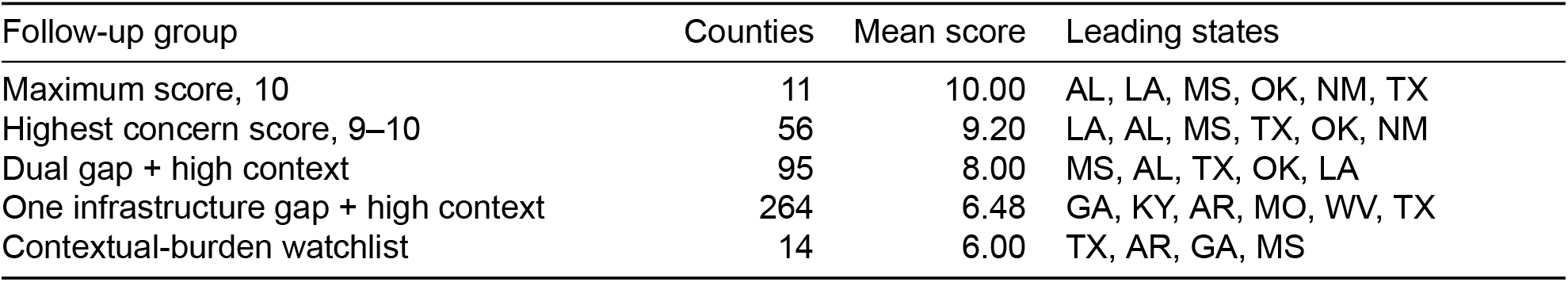
County Follow-Up Groups Identified by the Two-Tier Screening Framework.

**Supplemental Table S3.** Contextual-Burden Watchlist Counties. These 14 high-concern counties were driven by contextual burden without either direct infrastructure gap.

| County | State | Score |
| --- | --- | --- |
| Drew County | Arkansas | 6 |
| Jackson County | Arkansas | 6 |
| Crisp County | Georgia | 6 |
| Tift County | Georgia | 6 |
| Whitley County | Kentucky | 6 |
| Adair County | Missouri | 6 |
| Coahoma County | Mississippi | 6 |
| Washington County | Mississippi | 6 |
| Hertford County | North Carolina | 6 |
| McKinley County | New Mexico | 6 |
| Frio County | Texas | 6 |
| Nolan County | Texas | 6 |
| Starr County | Texas | 6 |
| San Juan County | Utah | 6 |

## Notes

### Competing Interest Statement

The authors have declared no competing interest.

